# Clinical Study Protocol of the ‘Biomarkers of Severity of COVID-19 Patients’ (BIOMARCOVID) Project

**DOI:** 10.64898/2026.06.16.26355763

**Authors:** Tuan-Anh Dinh, Corentin Leroy, Marion Brandolini-Bunlon, Sylvie Berthier, Candice Trocme, Nelle Varoquaux, Caroline Plazy, Antoine Vilotitch, Charles Terra, Bertrand Toussaint, Jean-Luc Bosson, Florence Castelli, Estelle Pujos-Guillot, Pauline Le Faouder, Justine Bertrand-Michel, Marion Le Marechal, Olivier Epaulard, Audrey Le Gouellec

## Abstract

**Introduction:** The coronavirus disease 2019 (COVID-19) pandemic has challenged health care systems worldwide, in certain areas exceeding hospital capacities and human resources. This has underscored the importance of having better tools to predict the outcome of potentially severe respiratory infections such as SARS-CoV-2. Predicting COVID-19 severity may allow physicians to better manage ICU beds and increase the chances of patient survival through appropriate management. During the toughest months of the pandemic, most physicians tried to identify patients that might develop severe forms based primarily on clinical features on admission (e.g., BMI, age). In this context, significant research has focused on identifying comorbidities, clinical manifestations, and routine blood biomarkers to predict disease severity. However, despite the demonstrated value of untargeted metabolomics in assessing severity, limited data exist on its use for identifying novel metabolite biomarkers that could improve both the sensitivity and specificity of outcome prediction. Our goal is to identify metabolite biomarkers that could enhance the predictive accuracy of standard medical biology data and clinical parameters.

**Methods and analysis:** This is a retrospective, observational, monocentric cohort study conducted at the Centre Hospitalier Universitaire Grenoble Alpes (CHUGA). The maximum number of eligible patients admitted for PCR-confirmed COVID-19 between March and December 2020 will be included. Severity outcome is defined using the WHO 10-category ordinal scale (mild: categories 4–5; severe: >5). Blood samples were collected within 48 hours of admission and analyzed for 62 routine blood tests and untargeted multiplatform LC-MS/MS metabolomics across four national platforms. Statistical analysis will include logistic regression with variable selection for the primary aim, and multi-block chemometric integration of clinical, biological, and metabolomics data as a secondary aim.

**Ethics and dissemination:** A study steering committee has been formed to ensure the accuracy of the collected data by thoroughly reviewing it prior to the data lock. All aspects of the study comply with ethical standards, including approval by the CHUGA institutional review board and adherence to CNIL Reference Methodology MR004 for the protection of participants’ rights, privacy, and confidentiality. This study is registered on the French Health Data Hub (number F20210218154851). Results will be disseminated through peer-reviewed publications, presentations at national and international scientific and clinical conferences, and reports shared with key healthcare system stakeholders.

**ARTICLE SUMMARY:** **Strengths and limitations of this study**

- Blood samples were collected within 48 hours of admission and before any severe symptom onset, aliquoted within 4 hours of collection and stored at −80°C, ensuring high pre-analytical quality.
- This study uses a multiplatform untargeted LC-MS/MS metabolomics approach across four complementary analytical platforms (CEA Saclay, INRAE Clermont, MetaToul, GEMELI), providing broad metabolome coverage.
- The integration of three heterogeneous data blocks (Metabolome, Biologicome, Clinicome) via multi-block chemometrics enables a systems-level view of COVID-19 severity.
- The sample size is limited by the complexity and cost of metabolomics analyses, and no formal sample size calculation was performed; findings should be considered exploratory and hypothesis-generating.
- As a monocentric, retrospective study conducted during a single epidemic wave (March–December 2020), generalizability may be limited by incomplete data across some routine blood tests and by differences in settings, subsequent variants, or treated populations.

## INTRODUCTION

The COVID-19 pandemic profoundly disrupted hospital operations, placing physicians worldwide in unprecedented circumstances. Patient stratification for those hospitalized with COVID-19 can be accomplished through clinical characteristics, ^1^·^2^ clinical symptomatology, ^3^ and routine blood biomarkers analyzed in medical biology laboratories. □However, the ability to predict disease severity using such data has proven to lack both specificity and sensitivity. This underscores the importance of ongoing research to identify more effective biomarkers for predicting disease progression.

At the Centre Hospitalier Universitaire Grenoble Alpes (CHUGA) in March 2020, when admitting patients without severe symptoms, predictions were primarily based on clinical features and emerging evidence from the literature, including age, sex, comorbidities (e.g., arterial hypertension, body mass index (BMI)^1^), and clinical symptoms.^3^ Nevertheless, due to the lack of sensitivity and specificity of these factors, an important “grey area” remains: physically fit and relatively young patients without known pre-existing conditions may develop severe COVID-19. Validated prognostic biomarkers include C-reactive protein (CRP) and IL-6,□as well as platelet count, N-terminal prohormone of brain natriuretic peptide (NT-proBNP), troponin I, lactate dehydrogenase (LDH), aspartate transaminase (AST), alanine transaminase (ALT), albumin, creatinine, urea, creatine kinase (CK), and white blood cell count (WBC).□□^1^□ However, the prognostic value of several other parameters remains insufficiently established, including agranular neutrophils (AGRAN), hemoglobin, red blood cell count, cholesterol, triglycerides, high-density lipoprotein (HDL), alkaline phosphatase (ALP), sodium, potassium, chloride, pH, red cell distribution width, gamma-glutamyl transferase (GGT), mean corpuscular hemoglobin concentration (MCHC), calcitriol (25-OHD), and total proteins.

Untargeted metabolomics offers significant advantages for biomarker discovery. It enables a comprehensive, hypothesis-free analysis of all metabolites detectable in biological samples, ^11^ making it well suited for identifying novel markers not captured by targeted methods. To date, untargeted metabolomics has been used primarily to study metabolic perturbations induced by SARS-CoV-2 infection, ^12^□^1^□ or to explore biomarkers of COVID-19 severity, ^1^□ with promising but preliminary results. Adding metabolite biomarkers identified by untargeted metabolomics to standard prognostic tests has the potential to improve accuracy, sensitivity, and specificity, and to reveal specific metabolic pathways involved in the response to infection.

This project is one of the few to combine prognostic biomarker discovery via untargeted metabolomics with routine blood test data in hospitalized COVID-19 patients and aims to provide a methodology adaptable to future emerging infectious diseases.

## METHODS AND ANALYSIS

### Study design and aims

BIOMARCOVID is a monocentric, retrospective cohort study of patients hospitalized for COVID-19 at CHUGA. The primary aim is to build a logistic regression model integrating clinical and biochemical parameters collected at hospitalization to predict disease severity outcome. Secondary aims are: (1) to identify differential metabolites and metabolic pathways between mild and severe outcomes using untargeted metabolomics; and (2) to perform an integrated multi-block analysis combining metabolomics, routine blood test, and clinical data. A biobank of clinical, biological, and metabolomic data will be constituted to support future sub-studies.

### Participants

#### Eligible participants must meet all inclusion criteria (Figure 1)

- Admitted to CHUGA for suspected COVID-19
- Confirmed positive PCR test for SARS-CoV-2

**Figure 1.**
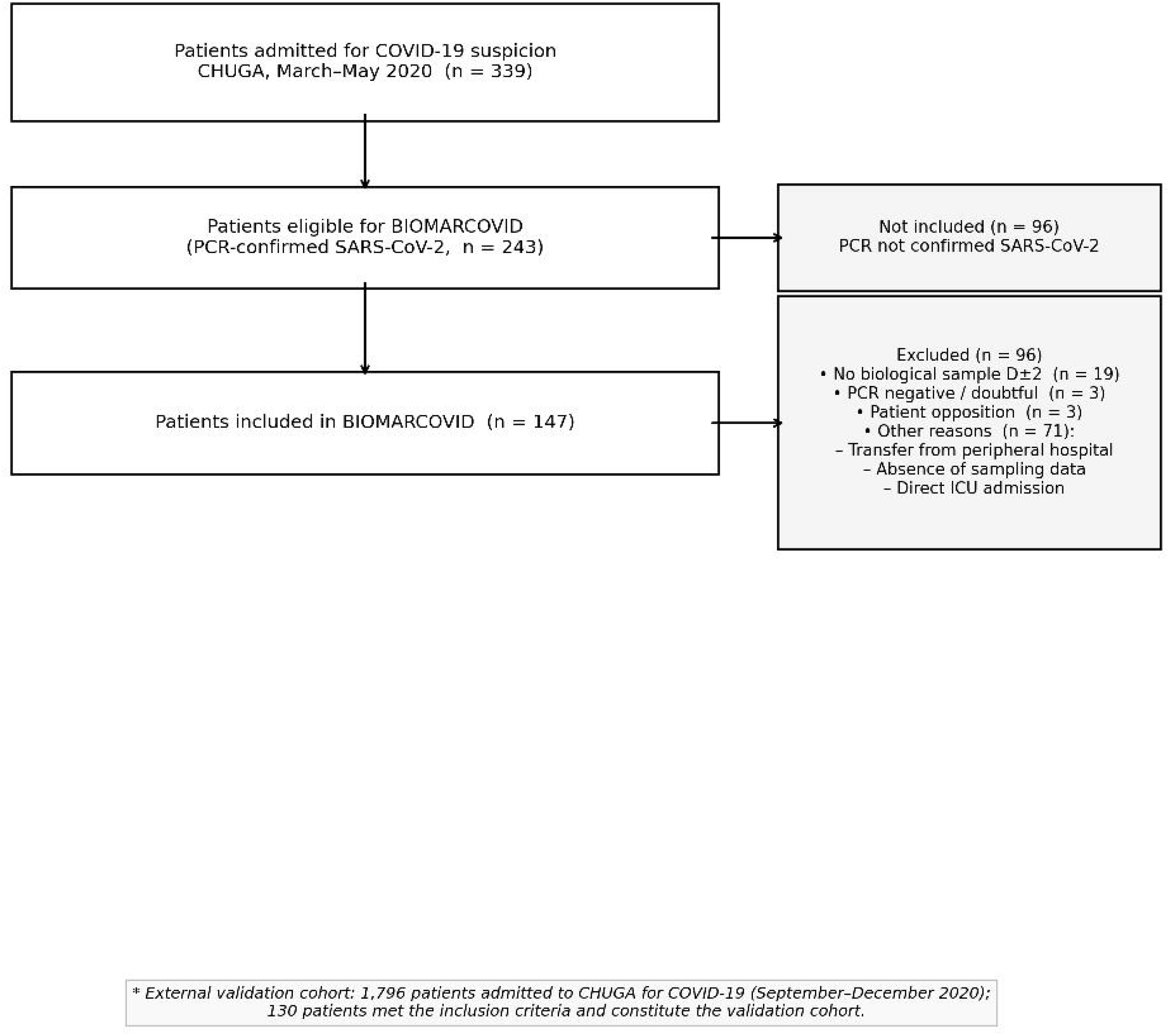
Flow diagram of participant selection (STROBE). Of 339 patients admitted to CHUGA for COVID-19 suspicion (March–May 2020), 243 were PCR-confirmed for SARS-CoV-2 and eligible for inclusion. A total of 147 patients were included in the BIOMARCOVID training cohort after applying exclusion criteria: absence of biological sampling within ±48 hours of admission (n=19); PCR-negative or doubtful result (n=3); patient opposition to research use of data (n=3); other reasons including transfer from a peripheral hospital, absence of sampling data, or direct ICU admission on day 1 (n=71). An external validation cohort was constituted from 1,796 patients admitted during the second wave (September–December 2020), of whom 130 met the inclusion criteria.

#### Exclusion criteria

- Age <18 years
- Transferred from another hospital
- Expressed opposition to research
- Admitted directly to the ICU on day 1
- Blood samples collected more than 48 hours after admission to CHUGA

### Data collection

#### Clinical evaluation (Clinicome)

Demographic data (age, sex, BMI), medical history (comorbidities including arterial hypertension, obesity, diabetes), and disease progression data (date of first symptoms, date of hospitalization, date of ICU admission and discharge, date of oxygen initiation, date of corticosteroid initiation, date of return home, live/death status) will be recorded for all patients. Involvement of ICU and infectious disease medical teams will support complete clinical data acquisition.

#### Patient-reported outcome measures

Patients will be classified retrospectively using the WHO 10-category ordinal progression scale: ^1^□ 0 (uninfected) to 10 (dead). For this study, patients with WHO scores of 4 or 5 (hospitalized, not requiring or requiring low-flow oxygen) are classified as mild COVID-19, and those with scores >5 as severe COVID-19.

#### Biological samples (Biologicome)

Sixty-two routine blood tests will be included (see online supplemental data 1). These tests were performed according to CHUGA standard procedures upon patient admission. Pre-analytical steps and pricing are described in CHUGA’s biological catalogue (http://www.monkiosquesante.org/KS_EXP/LivretBio). For routine blood tests, samples were analysed prospectively as part of standard clinical care; no randomisation was applicable given the retrospective nature of the study.

#### Metabolomics (Metabolome)

Residual EDTA plasma samples, initially collected for IL-6 assays, will be repurposed for untargeted metabolomics analyses using multiplatform LC-MS/MS at four national platforms (CEA Saclay, INRAE Clermont, MetaToul, GEMELI). Samples are processed within 4 hours of collection, aliquoted, and stored at −80°C until analysis. Metabolites are extracted from plasma proteins by methanol precipitation prior to analysis. Clinical and biological data are pseudonymised and stored in a secure database with access restricted to the study team, in compliance with CNIL MR004 requirements. Sample injection order was randomised across all four analytical platforms in accordance with mQACC quality assurance recommendations, to minimise systematic analytical drift and batch effects.

### Metabolomics data processing

Raw files will be converted to mzXML format using MSConvert. Automatic peak detection and integration will be performed using the XCMS software package. ^2^□ The resulting data matrix contains m/z values, retention times, and peak areas (arbitrary units). XCMS features will be filtered based on: (i) correlation between QC dilution factors and peak areas (r >0.7); (ii) repeatability (coefficient of variation of QC peak areas <30%); and (iii) biological-to-blank sample peak area ratio >3. If required, LOESS normalization will be applied to correct analytical drift. Additionally, metabolomics features with more than 30% missing values across samples will be excluded from analysis.

### Statistical analysis

#### General considerations

This is a pilot study; no formal sample size calculation was performed. All eligible patients admitted to CHUGA during the study period will be included. Quantitative variables will be described using median and interquartile range; categorical variables using count and percentage. Complete case analysis will be used for the primary outcome. Additional targeted assays were performed to reduce missing data across routine blood tests. Statistical significance is set at α=0.05.

#### Primary analysis: clinical and biochemical factors

The primary aim is to build a logistic regression model with LASSO variable selection, using clinical and routine biochemical parameters at admission to predict severity outcome. Model performance will be evaluated using accuracy, sensitivity, specificity, positive predictive value, and negative predictive value, as well as the cost of biological parameters (Figure 2). Routine blood test variables with more than 80% missing values across samples will be excluded from analysis. Blinding was applied where appropriate. Unsupervised analyses (PCA, multi-block exploratory analyses) were performed without knowledge of severity group allocation, allowing unbiased exploration of data structure. For supervised analyses (logistic regression with LASSO variable selection, discriminant analyses), severity class labels were provided to the model as the outcome variable, as required by the analytical approach.

**Figure 2.**
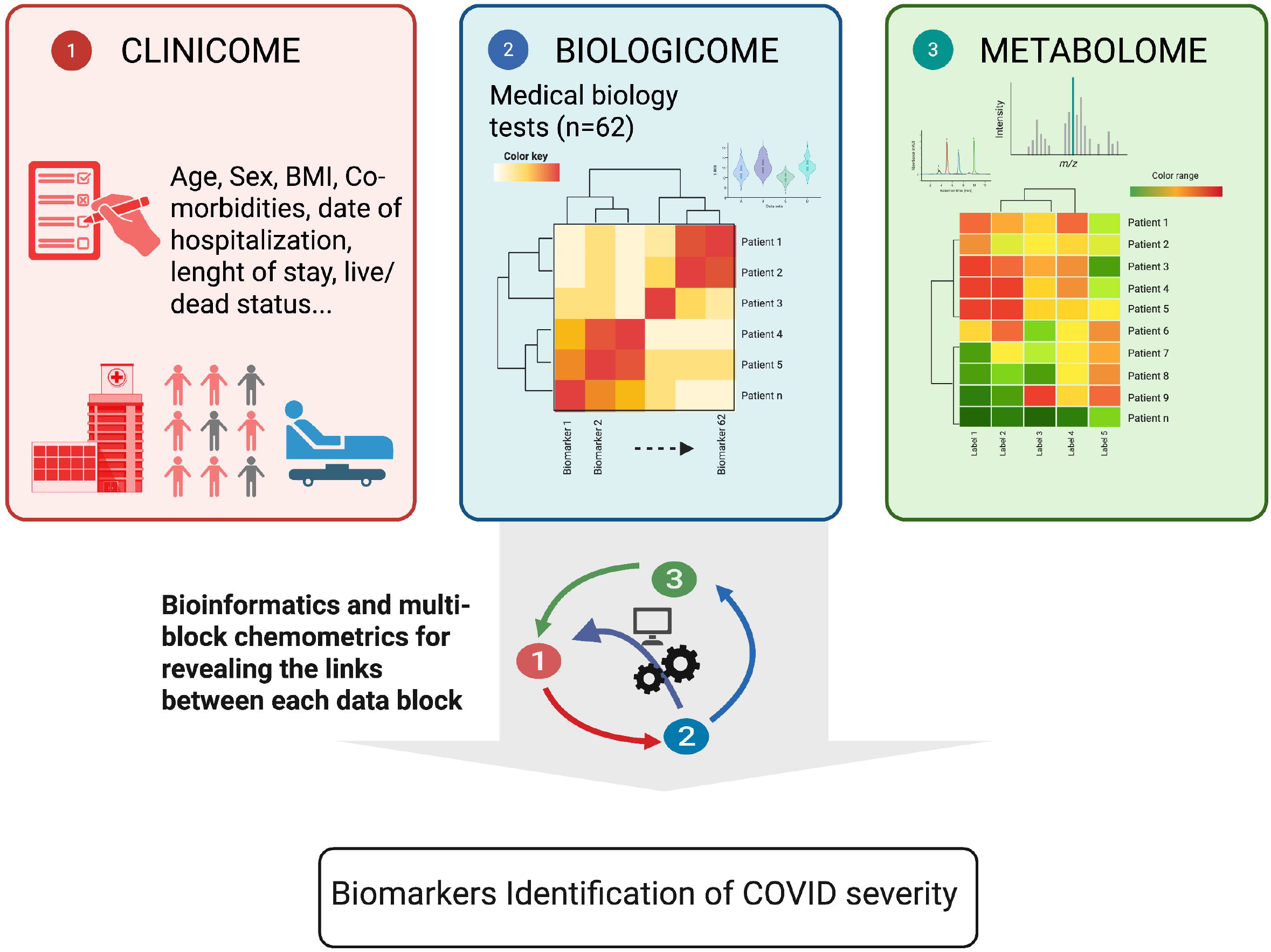
Study principle. Three data blocks, Metabolome (untargeted LC-MS/MS), Biologicome (routine laboratory tests), and Clinicome (clinical characteristics), are analyzed individually, then integrated using multi-block chemometrics to predict COVID-19 severity outcome. Created using BioRender (https://BioRender.com).

#### Secondary analysis: metabolomics and lipidomics biomarkers

Each routine laboratory test and metabolite will be tested individually and in combination for their ability to predict severe COVID-19. Principal component analysis (PCA) will be performed on each data matrix (Biologicome and Metabolome) to explore variance and identify co-factors. Routine blood variables will be normalized by their standard deviation; metabolomics variables will be log-transformed and Pareto-scaled. Logistic regressions with sex, age, and obesity as cofactors will be used to identify predictive variables, with p-values adjusted by the Benjamini-Hochberg method. Significant variables will be decorrelated, and a reduced logistic regression model will be selected by AIC and validated by 10-fold cross-validation. No pre-planned subgroup or sensitivity analyses are planned beyond those described above.

#### Multi-block analysis

The study is structured around three data blocks: Metabolome (untargeted LC-MS/MS data), Biologicome (62 routine blood tests), and Clinicome (clinical characteristics and symptoms). Single-block analyses will first be performed to compute block-specific scores and loadings. A multi-block modelling approach will then be applied to integrate all data sources, provide a global view of their combined information, assess the relative contribution of each block, and identify cross-block variable associations. ^21^·^22^

## ETHICS AND DISSEMINATION

This study is conducted in accordance with International Conference on Harmonization Good Clinical Practice guidelines. It is a non-interventional, monocentric, retrospective study involving data and samples from human participants, conducted at CHUGA in compliance with French regulations. The principal investigator (Prof. Olivier Epaulard) has signed a commitment to Reference Methodology No. 004 (MR004) issued by the French Data Protection Authority (CNIL). The study is registered on the French Health Data Hub under number F20210218154851. All participants were informed; none expressed opposition. Written informed consent was not required under French national legislation and institutional guidelines. This protocol will be made available upon publication via BMJ Open. Any substantial amendments to this protocol will be submitted to the CHUGA IRB and the CNIL for approval prior to implementation and communicated to all co-investigators.

### Patient and public involvement

Patients were not involved in the design, conduct, or reporting of this study. This was a retrospective study using residual biological samples and routine clinical data collected during standard care. Results will be disseminated to the public through lay summaries and conference presentations.

### Data availability

The statistical code and analytical pipeline supporting this study are available from the Zenodo repository (DOI: 10.5281/zenodo.20558738). The metabolomics data and metadata are available via MassIVE upon reasonable request, in compliance with the General Data Protection Regulation.

A steering committee comprising intensive care physicians, infectious disease specialists, medical biologists, and statisticians will validate data accuracy prior to data lock and oversee sub-study project selection. Results will be published in peer-reviewed journals and presented at national and international scientific and clinical conferences.

## DISCUSSION

Numerous studies have investigated blood biomarkers and clinical risk factors to predict the severity and mortality of COVID-19.^1^□^1^□ Elevated systemic cytokine levels, immune activation, and organ injury markers have been documented, driven in part by the angiotensin-converting enzyme 2 (ACE2) receptor, which is expressed in multiple organs and serves as the functional entry point for SARS-CoV-2.^23^

Understanding ACE2-mediated pathophysiology is critical because many factors — including age, sex, ethnicity, comorbidities, and medication — influence both ACE2 expression and COVID-19 severity. However, ACE2-expressing organs do not equally participate in COVID-19 pathophysiology, implying that additional mechanisms contribute to tissue damage and disease severity. ^23^

BIOMARCOVID addresses this gap by coupling 62 routine blood tests with untargeted multiplatform metabolomics in a well-characterized cohort of hospitalized patients. The use of plasma samples collected within 48 hours of admission — before severe symptom onset — is a key methodological strength, as it ensures that biomarkers reflect early pathophysiological processes rather than late-stage complications.

Metabolomic profiling has the potential to differentiate mild from severe SARS-CoV-2 infection and to identify metabolic pathways dysregulated in early disease. Combined with routine blood test data and clinical parameters through a multi-block analytical framework, this approach may enable more precise patient stratification, better anticipation of ICU bed requirements, and identification of early therapeutic targets. ^11^ Importantly, the methodology developed here is designed to be transferable to other emerging respiratory infections.

The main limitations of this study are its monocentric, retrospective design; the absence of a formal sample size calculation; incomplete data for some routine tests; and the potential influence of self-medication or variable time from symptom onset to admission on metabolomic profiles. These limitations are addressed in part by the multi-platform metabolomics approach, data imputation strategies, and the prospective constitution of a biobank for future validation studies.

## Supporting information

https://zenodo.org/records/20715711/files/BIOMARCOVID_SPIRIT2025_checklist_final.docx?download=1

## AUTHOR CONTRIBUTIONS

ALG and OE conceived and designed the clinical study. SB, PLF, CT and TD conducted experiments. ALG, EP, FC, JBM, MB, CL, AV and OE will analyze the data data. TD and ALG wrote the manuscript. All authors read, reviewed, and approved the final manuscript.

## LICENCE STATEMENT A

This article is submitted under the Creative Commons Attribution licence (CC BY). This licence permits reuse, distribution, and reproduction in any medium, provided the original work is properly cited.

## ETHICS APPROVAL AND CONSENT TO PARTICIPATE

This research was approved by the CHUGA institutional review board and authorized following filing with the CNIL under French procedure for a monocentric study (MR004 study). All patients were informed and none expressed opposition to research use of their data; written consent was not required under national legislation.

## ACKNOWLEDGEMENTS

We warmly thank Z. Baidi for assistance with this research.

## REFERENCES

1. Gao Y, Ding M, Dong X, et al. Risk factors for severe and critically ill COVID-19 patients: a review. Allergy 2021;76:428–55. doi:10.1111/all.14657

2. Hu B, Guo H, Zhou P, et al. Characteristics of SARS-CoV-2 and COVID-19. Nat Rev Microbiol 2021;19:141–54. doi:10.1038/s41579-020-00459-7

3. Ye Z, Zhang Y, Wang Y, et al. Chest CT manifestations of new coronavirus disease 2019 (COVID-19): a pictorial review. Eur Radiol 2020;30:4381–9. doi:10.1007/s00330-020-06801-0

4. Gallo Marin B, Aghagoli G, Lavine K, et al. Predictors of COVID-19 severity: a literature review. Rev Med Virol 2021;31:1–10. doi:10.1002/rmv.2146

5. Izcovich A, Ragusa MA, Tortosa F, et al. Prognostic factors for severity and mortality in patients infected with COVID-19: a systematic review. PLoS One 2020;15:e0241955. doi:10.1371/journal.pone.0241955

6. Liu F, Li L, Xu M, et al. Prognostic value of interleukin-6, C-reactive protein, and procalcitonin in patients with COVID-19. J Clin Virol 2020;127:104370. doi:10.1016/j.jcv.2020.104370

7. Karimi A, Shobeiri P, Kulasinghe A, et al. Novel systemic inflammation markers to predict COVID-19 prognosis. Front Immunol 2021;12:741061. doi:10.3389/fimmu.2021.741061

8. Terpos E, Ntanasis-Stathopoulos I, Elalamy I, et al. Hematological findings and complications of COVID-19. Am J Hematol 2020;95:834–47. doi:10.1002/ajh.25829

9. Battaglini D, Lopes-Pacheco M, Castro-Faria-Neto HC, et al. Laboratory biomarkers for diagnosis and prognosis in COVID-19. Front Immunol 2022;13:857573. doi:10.3389/fimmu.2022.857573

10. Tjendra Y, Al Mana AF, Espejo AP, et al. Predicting disease severity and outcome in COVID-19 patients: a review of multiple biomarkers. Arch Pathol Lab Med 2020;144:1465–74. doi:10.5858/arpa.2020-0471-SA

11. Le Gouellec A, Plazy C, Toussaint B. What clinical metabolomics will bring to the medicine of tomorrow. Front Anal Sci 2023;3. doi:10.3389/frans.2023.1142606

12. Bi X, Liu W, Ding X, et al. Proteomic and metabolomic profiling of urine uncovers immune responses in patients with COVID-19. Cell Rep 2022;38:110271. doi:10.1016/j.celrep.2021.110271

13. Grassin-Delyle S, Roquencourt C, Moine P, et al. Metabolomics of exhaled breath in critically ill COVID-19 patients: a pilot study. EBioMedicine 2021;63:103154. doi:10.1016/j.ebiom.2020.103154

14. Mussap M, Fanos V. Could metabolomics drive the fate of COVID-19 pandemic? Clin Chem Lab Med 2021;59:1891–905. doi:10.1515/cclm-2021-0414

15. Shen B, Yi X, Sun Y, et al. Proteomic and metabolomic characterization of COVID-19 patient sera. Cell 2020;182:59–72. doi:10.1016/j.cell.2020.05.032

16. Song JW, Lam SM, Fan X, et al. Omics-driven systems interrogation of metabolic dysregulation in COVID-19 pathogenesis. Cell Metab 2020;32:188–202. doi:10.1016/j.cmet.2020.06.016

17. Wu D, Shu T, Yang X, et al. Plasma metabolomic and lipidomic alterations associated with COVID-19. Natl Sci Rev 2020;7:1157–68. doi:10.1093/nsr/nwaa086

18. Hasan MR, Suleiman M, Pérez-López A. Metabolomics in the diagnosis and prognosis of COVID-19. Front Genet 2021;12:721556. doi:10.3389/fgene.2021.721556

19. WHO Working Group on the Clinical Characterisation and Management of COVID-19 infection. A minimal common outcome measure set for COVID-19 clinical research. Lancet Infect Dis 2020;20:e192–7. doi:10.1016/S1473-3099(20)30483-7

20. Giacomoni F, Le Corguillé G, Monsoor M, et al. Workflow4Metabolomics: a collaborative research infrastructure for computational metabolomics. Bioinformatics 2015;31:1493–5. doi:10.1093/bioinformatics/btu813

21. Boccard J, Rudaz S. Harnessing the complexity of metabolomic data with chemometrics. J Chemom 2014;28:1–9. doi:10.1002/cem.2567

22. Boccard J, Rutledge DN. A consensus orthogonal partial least squares discriminant analysis (OPLS-DA) strategy for multiblock Omics data fusion. Anal Chim Acta 2013;769:30–9. doi:10.1016/j.aca.2013.01.022

23. Bourgonje AR, Abdulle AE, Timens W, et al. Angiotensin-converting enzyme 2 (ACE2), SARS-CoV-2 and the pathophysiology of coronavirus disease 2019 (COVID-19). J Pathol 2020;251:228–48. doi:10.1002/path.5471

