## Supplementary material for "Clinical Study Protocol of the ‘Biomarkers of Severity of COVID-19 Patients’ (BIOMARCOVID) Project": https://zenodo.org/records/20715711/files/BIOMARCOVID_SPIRIT2025_checklist_final.docx?download=1

**SPIRIT 2025 Checklist — BIOMARCOVID Protocol**

*Biomarkers of Severity of COVID-19 Patients | BMJ Open | Version n°3.0 du 23.05.2024*

**Note:** *SPIRIT 2025 was developed for randomised trials. BIOMARCOVID is a retrospective observational cohort study. Items on randomisation and blinding are N/A. All 34 items addressed.*

**✅ Present N/A Not applicable**

**1. ADMINISTRATIVE INFORMATION**

| **No** | **SPIRIT 2025 Item** | **Status** | **Page in manuscript (v2, 3 June 2026)** |
| --- | --- | --- | --- |
| 1a | Title stating the trial design, population, and interventions, identified as a protocol | **✅ Present** | Page 1 — "Clinical Study Protocol… BIOMARCOVID Project" |
| 1b | Structured summary including WHO Trial Registration Data Set items | **✅ Present** | Page 2 — Abstract: Introduction / Methods and analysis / Ethics and dissemination |
| 2 | Protocol version date and identifier | **✅ Present** | Page 1 — "Version n°3.0 du 23.05.2024" |
| 3a | Names, affiliations, and roles of contributors | **✅ Present** | Page 1 (authors + affiliations) + Page 7 (Contributors section) |
| 3b | Name and contact information for the trial sponsor | **✅ Present** | Page 1 — "Sponsor: CHU Grenoble Alpes, CS 10217, 38043 Grenoble Cedex 9, France" |
| 3c | Role of sponsor and funders in design, conduct, analysis, and reporting | **✅ Present** | Page 8 — "The funders had no role in study design, data collection, analysis, decision to publish, or preparation of the manuscript." |
| 3d | Composition and roles of steering committee and other oversight groups | **✅ Present** | Page 6 — Steering committee described in Ethics and Dissemination section |

**2. OPEN SCIENCE**

| **No** | **SPIRIT 2025 Item** | **Status** | **Page in manuscript (v2, 3 June 2026)** |
| --- | --- | --- | --- |
| 4 | Trial registry name, identifying number (URL), and date of registration | **✅ Present** | Page 6 — French Health Data Hub, registration number F20210218154851 |
| 5 | Where the trial protocol and statistical analysis plan can be accessed | **✅ Present** | Page 6 — "This protocol will be made available upon publication via BMJ Open." |
| 6 | Data sharing: where de-identified participant data will be accessible | **✅ Present** | Page 6 — MassIVE upon reasonable request, in compliance with GDPR |
| 7a | Sources of funding and other support | **✅ Present** | Page 7–8 — VLM, AGL (RF20230503289), ANR-15-IDEX-02, FINOVI, Fondation UGA, GEMELI-GExiM platform |
| 7b | Financial and other conflicts of interest | **✅ Present** | Page 8 — "All authors declare no conflict of interest" |
| 8 | Dissemination policy: plans to communicate results to participants and the public | **✅ Present** | Page 6 — Patient and public involvement statement + dissemination plan |

**3. INTRODUCTION**

| **No** | **SPIRIT 2025 Item** | **Status** | **Page in manuscript (v2, 3 June 2026)** |
| --- | --- | --- | --- |
| 9a | Scientific background and rationale, including summary of relevant studies | **✅ Present** | Pages 3–6 — Introduction + Discussion |
| 9b | Explanation for choice of comparator | **N/A** | Observational study — no experimental intervention or comparator |
| 10 | Specific objectives | **✅ Present** | Page 2 (Abstract: Methods and analysis) + Page 4 (Study design and aims) |

**4. METHODS: PATIENT AND PUBLIC INVOLVEMENT, TRIAL DESIGN**

| **No** | **SPIRIT 2025 Item** | **Status** | **Page in manuscript (v2, 3 June 2026)** |
| --- | --- | --- | --- |
| 11 | Details of patient or public involvement in design, conduct, and reporting | **✅ Present** | Page 6 — Patient and public involvement statement |
| 12 | Description of trial design including type, allocation ratio, and framework | **✅ Present** | Page 2 (Abstract) + Page 4 — "Retrospective, observational, monocentric cohort study" |

**5. METHODS: PARTICIPANTS, INTERVENTIONS, AND OUTCOMES**

| **No** | **SPIRIT 2025 Item** | **Status** | **Page in manuscript (v2, 3 June 2026)** |
| --- | --- | --- | --- |
| 13 | Settings and locations where study will be conducted | **✅ Present** | Page 2 (Abstract) + Page 4 — CHUGA, Grenoble, France, March–December 2020 |
| 14a | Eligibility criteria for participants | **✅ Present** | Page 4 — Inclusion and exclusion criteria listed |
| 14b | Eligibility criteria for sites and individuals delivering interventions | **N/A** | Single-centre observational study |
| 15a | Intervention and comparator with sufficient detail to allow replication | **N/A** | Observational study — no experimental intervention |
| 15b | Criteria for discontinuing or modifying allocated intervention | **N/A** | No intervention |
| 15c | Strategies to improve adherence to intervention protocols | **N/A** | No intervention |
| 15d | Concomitant care permitted or prohibited | **N/A** | Observational study — concomitant care not controlled |
| 16 | Primary and secondary outcomes: measurement variable, analysis metric, time point | **✅ Present** | Page 2 (Abstract) + Page 4 (Study design) + Page 4 (WHO scale, mild vs severe during hospitalisation) |
| 17 | How harms are defined and will be assessed | **N/A** | Observational study using residual samples and routine data — no experimental harms |
| 18 | Time schedule of enrolment, interventions, and assessments | **✅ Present** | Page 7 — Figure 1 (CONSORT flowchart of participant selection and exclusion) |
| 19 | How sample size was determined | **✅ Present** | Page 5 — Pilot study; all eligible patients included; no formal sample size calculation; justification provided |
| 20 | Strategies for achieving adequate participant enrolment | **✅ Present** | Page 5 — All eligible patients admitted to CHUGA between March and December 2020 |

**6. METHODS: ASSIGNMENT OF INTERVENTIONS (N/A — OBSERVATIONAL STUDY)**

| **No** | **SPIRIT 2025 Item** | **Status** | **Page in manuscript (v2, 3 June 2026)** |
| --- | --- | --- | --- |
| 21a | Random allocation sequence generation | **N/A** | Non-randomised observational study |
| 21b | Type of randomisation and stratification | **N/A** | Non-randomised observational study |
| 22 | Allocation concealment mechanism | **N/A** | Non-randomised observational study |
| 23 | Implementation of allocation sequence | **N/A** | Non-randomised observational study |
| 24a | Who will be blinded after assignment | **N/A** | Non-randomised observational study |
| 24b | How blinding will be achieved | **N/A** | Non-randomised observational study |
| 24c | Circumstances permitting unblinding | **N/A** | Non-randomised observational study |

**7. METHODS: DATA COLLECTION, MANAGEMENT, AND ANALYSIS**

| **No** | **SPIRIT 2025 Item** | **Status** | **Page in manuscript (v2, 3 June 2026)** |
| --- | --- | --- | --- |
| 25a | Plans for data collection, instruments, reliability and validity | **✅ Present** | Pages 4–5 — Clinicome (Page 4), Biologicome (Pages 4–5), Metabolome (Page 5). Reference to CHUGA biological catalogue and online supplemental data 1. |
| 25b | Plans to promote participant retention and complete follow-up | **✅ Present** | Retrospective study — all data collected at admission. Loss to follow-up not applicable for the primary endpoint. |
| 26 | Data management: entry, coding, security, and storage | **✅ Present** | Page 5 — "Clinical and biological data are pseudonymised and stored in a secure database with access restricted to the study team, in compliance with CNIL MR004 requirements." |
| 27a | Statistical methods for primary and secondary outcomes | **✅ Present** | Pages 5–6 — Logistic regression with LASSO (Page 5), PCA + Benjamini-Hochberg correction (Pages 5–6), multi-block chemometrics (Page 6) |
| 27b | Definition of analysis population | **✅ Present** | Page 5 — "All eligible patients admitted to CHUGA during the study period will be included." |
| 27c | How missing data will be handled | **✅ Present** | Page 5 — "Complete case analysis will be used for the primary outcome. Additional targeted assays were performed to reduce missing data across routine blood tests." |
| 27d | Methods for additional analyses (subgroup, sensitivity) | **✅ Present** | Page 6 — "No pre-planned subgroup or sensitivity analyses are planned beyond those described above." |

**8. METHODS: MONITORING**

| **No** | **SPIRIT 2025 Item** | **Status** | **Page in manuscript (v2, 3 June 2026)** |
| --- | --- | --- | --- |
| 28a | Data monitoring committee composition, role, and independence | **✅ Present** | Page 6 — Steering committee described. Given the retrospective observational design, a formal independent DMC is not required. |
| 28b | Interim analyses and stopping guidelines | **N/A** | Retrospective study — no interim analysis applicable |
| 29 | Frequency and procedures for monitoring trial conduct | **✅ Present** | Page 6 — Steering committee validates data accuracy prior to data lock |

**9. ETHICS**

| **No** | **SPIRIT 2025 Item** | **Status** | **Page in manuscript (v2, 3 June 2026)** |
| --- | --- | --- | --- |
| 30 | Plans for seeking research ethics committee / IRB approval | **✅ Present** | Page 6 (Ethics and Dissemination) + Page 8 (Ethics Approval section) — CHUGA IRB approved, CNIL MR004, Health Data Hub F20210218154851 |
| 31 | Plans for communicating important protocol modifications | **✅ Present** | Page 6 — "Any substantial amendments to this protocol will be submitted to the CHUGA IRB and the CNIL for approval prior to implementation and communicated to all co-investigators." |
| 32a | Who will obtain informed consent and how | **✅ Present** | Page 6 + Page 8 — All participants informed; none expressed opposition. Written consent not required under French national legislation. |
| 32b | Additional consent for ancillary studies / biological specimens | **✅ Present** | Page 4 — Biobank for future sub-studies constituted; participants informed per CNIL MR004 framework. |
| 33 | Confidentiality: how personal information will be protected | **✅ Present** | Page 5 (pseudonymisation explicitly stated) + Page 6 (CNIL MR004 compliance) |
| 34 | Provisions for post-trial care; compensation for harms | **N/A** | Observational study using residual samples and routine clinical data. No experimental intervention; no harm from study participation. |
